# Highly-specific memory B cells generation after the 2^nd^ dose of BNT162b2 vaccine compensate for the decline of serum antibodies and absence of mucosal IgA

**DOI:** 10.1101/2021.06.08.21258284

**Authors:** Eva Piano Mortari, Cristina Russo, Maria Rosaria Vinci, Sara Terreri, Ane Fernandez Salinas, Livia Piccioni, Claudia Alteri, Luna Colagrossi, Luana Coltella, Stefania Ranno, Giulia Linardos, Marilena Agosta, Christian Albano, Chiara Agrati, Concetta Castilletti, Silvia Meschi, Paolo Romania, Giuseppe Roscilli, Emiliano Pavoni, Vincenzo Camisa, Annapaola Santoro, Rita Brugaletta, Nicola Magnavita, Alessandra Ruggiero, Nicola Cotugno, Donato Amodio, Marta Luisa Ciofi Degli Atti, Daniela Giorgio, Nicoletta Russo, Guglielmo Salvatori, Tiziana Corsetti, Franco Locatelli, Carlo Federico Perno, Salvatore Zaffina, Rita Carsetti

**Affiliations:** Diagnostic Immunology Research Unit, Multimodal Medicine Research Area, Bambino Gesù Children’s Hospital, IRCCS; Viale di San Paolo,15, Rome, Italy.; Microbiology and Diagnostic Immunology Unit, Bambino Gesù Children’s Hospital, IRCCS; Piazza Sant’Onofrio, 4, Rome, Italy.; Occupational Medicine/Health Technology Assessment and Safety Research Unit, Clinical-Technological Innovations Research Area, Bambino Gesù Children’s Hospital, IRCCS, Viale di San Paolo, 15,Rome, Italy.; Health Directorate, Bambino Gesù Children’s Hospital, IRCCS; Piazza Sant’Onofrio, 4, Rome, Italy.; Department of Molecular Medicine, Sapienza University of Rome; Viale dell’Università, 37 Rome, Italy.; Department of Oncology and Hemato-Oncology, University of Milan, Via festa del Perdono,7,Milan, Italy.; National Institute for Infectious Diseases Lazzaro Spallanzani, IRCCS; Via Portuense, 2, Rome, Italy.; Takis s.r.l.; Via di Castel Romano, 100, Rome, Italy.; Post-Graduate School of Occupational Health, Section of Occupational Medicine and Labor Law, University Cattolica del Sacro Cuore; Largo Francesco Vito, 1, Rome, Italy; Department of Woman, Child & Public Health, Fondazione Policlinico Universitario A. Gemelli IRCCS; Via della Pineta Sacchetti, 217, Rome, Italy; Clinical and Research Unit of Clinical Immunology and Vaccinology, Bambino Gesù Children’s Hospital, IRCCS; Piazza Sant’Onofrio, 4, Rome, Italy.; Clinical Pathways and Epidemiology Unit, Bambino Gesù Children’s Hospital, IRCCS; Piazza Sant’Onofrio, 4, Rome, Italy.; Neonatal Intensive Care Unit and Human Milk Bank, Department of Neonatology, Bambino Gesù Children’s Hospital, IRCSS; Piazza Sant’Onofrio, 4, Rome, Italy.; Hospital Pharmacy Unit, Bambino Gesù Children’s Hospital, IRCCS; Piazza Sant’Onofrio, 4, Rome, Italy.; Department of Hematology/Oncology, Bambino Gesù Children’s Hospital, IRCCS; Piazza Sant’Onofrio, 4, Rome, Italy.; Department of Pediatrics, Sapienza, University of Rome; Viale dell’Università, 37, Rome, Italy.

## Abstract

Specific memory B cells and antibodies are reliable read-out of vaccine efficacy. We analyzed these biomarkers after one and two doses of BNT162b2 vaccine. The second dose significantly increases the level of highly-specific memory B cells and antibodies. Two months after the second dose, specific antibody levels decline, but highly specific memory B cells continue to increase thus predicting a sustained protection from COVID-19.

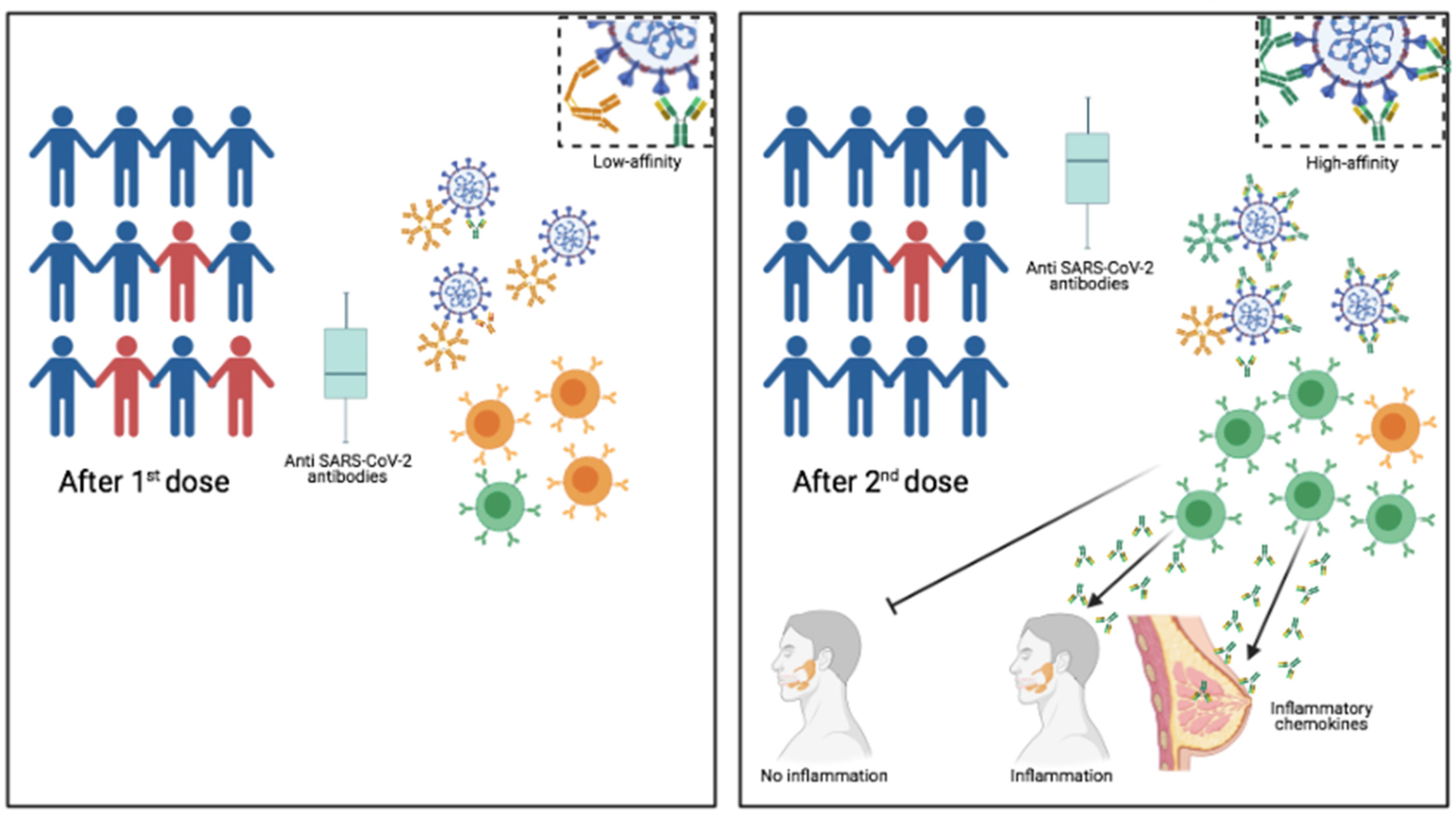
Graphical Abstract

---

We show that although mucosal IgA is not induced by the vaccination, memory B cells migrate in response to inflammation and secrete IgA at mucosal sites.

We show that first vaccine dose may lead to an insufficient number of highly specific memory B cells and low concentration of serum antibodies thus leaving vaccinees without the immune robustness needed to ensure viral elimination and herd immunity. We also clarify that the reduction of serum antibodies does not diminish the force and duration of the immune protection induced by vaccination. The vaccine does not induce sterile immunity. Infection after vaccination may be caused by the lack of local preventive immunity because of the absence of mucosal IgA.

The SARS-CoV-2 pandemic continues spreading and causing COVID-19 disease and death. Hospitals are under extreme pressure and the economic damage is unprecedented. Vaccination is the safest and most effective tool to achieve a protective response in most individuals. Effective vaccines against SARS-CoV-2 have been rapidly developed and are being administered with the aim of preventing COVID-19, stopping viral circulation and terminating the pandemic. Phase III trials have demonstrated the high efficacy of two-dose regimens^1–3^, but because of the low number of vaccine doses available in Europe, it has been suggested that initially only the first dose may be administered to the whole population in the emergency phase. The first dose may be able to prevent the most severe forms of disease, thus avoiding hospitalization and death^4^. According to current epidemiological and political opinions, the second dose may be delayed. Concerns have been raised that changing the immunization schedule proved to be effective by the authorization studies, because of results obtained in the real world on the effectiveness of the first dose ^5^ and on the consideration that incomplete immunity may favour the emergency of viral variants^6^. From the epidemiological point of view, it has been predicted that a one-dose regimen will foster antigenic evolution and weaken the protective potentials of the available vaccines and of those still in development, all based on the sequence of a virus that may have changed under the selective pressure of an immune response unable to completely clear the virus^7^. Several Variants of concern (VoC) have been identified and have spread throughout the world. New variants are continuously generated^8^.

Another still open question is the duration of protective immunity after vaccination. Most attention has been concentrated on the level of specific antibodies, which increase in response to the vaccine, but decline after a few months. The reduction of specific antibody levels generates considerable anxiety in the population, because it is perceived as incipient loss of protection.

Antibody decline is, however, a normal feature of the response to every vaccination^9^. Our most important protection from reinfection is represented by the synergistic action of memory B cells (MBCs) and memory plasma cells. Specific antibodies present in the serum are continuously secreted by memory plasma cells and face the pathogen at its first entry^10^. At the same time, MBCs migrate in response to chemokines to the inflamed tissues and reinforce protection by locally secreting antibodies. If the antibodies produced by MBCs and plasma cells have a high affinity for the pathogen this is immediately eliminated^9^. T cells play a fundamental role by helping B cells to produce high-affinity antibodies and memory B cells^11, 12^ and, most importantly, by eliminating virus infected cells^13^.

From the point of view of B-cell immunology, in order to support public health decisions and measures, the most important issues to be addressed are: 1. the affinity of MBCs and plasma cells generated after the 1^st^ and 2^nd^ vaccine dose; 2. the longevity of vaccine-induced immune memory; 3. the ability of the vaccine to induce sterilizing immunity, thus preventing not only COVID-19, but also infection.

We addressed these three points in a study on the B-cell response to the first and second dose of BNT162b2 vaccine and a further evaluation of the established protection three months after.

We report the effects of the two-doses regimen of BNT162b2 vaccine in a population of Health Care Workers (HCWs) of the Bambino Gesù Children Hospital IRCCS.

In 108 HCWs we measured specific MBCs by ELISpot, flow-cytometry and serum antibodies at different time points, before and after first and second dose and three months later. We found that one single dose results in immune memory lacking the precision indispensable to immediately clear the virus that is the basis for vaccine effectiveness. The second dose increases the frequency of vaccine responders and specific IgG levels and generates highly specific MBCs. Three months later, serum antibodies significantly decline, but, in contrast, MBCs increase in frequency and affinity. Most importantly, the vaccine triggers a serological IgA response, but does not generate mucosal IgA. The lack of specific IgA strategically located at the virus site of entrance explains why the vaccine does not induce sterilizing immunity.

## Results

### Vaccination of HCWs

We included in our study 108 HCWs (Table 1) who had never experienced SARS-CoV-2 infection before, as demonstrated by molecular (Allplex2019-ncov, Seegene) and antibody assays (Elecsys®Anti-N, Roche) (data not shown).

**Table 1:** Characteristics of the study population.

| Study Group |  |
| --- | --- |
| Number of HCW | 108 |
| Age (mean ± SD) | 46.95 ± 11.35 |
| Sex (M/F) | 31/77 |

The BNT162b2 vaccine was administered as prescribed, in two doses, 21 days apart. Serum and blood samples were collected at time 0 (T0), before the first dose; seven days after the first dose (T7); on the day of the second dose (T21), seven days after the second dose (T28) and 3 months after the first dose (T90) (Fig. 1).

**Fig. 1.**
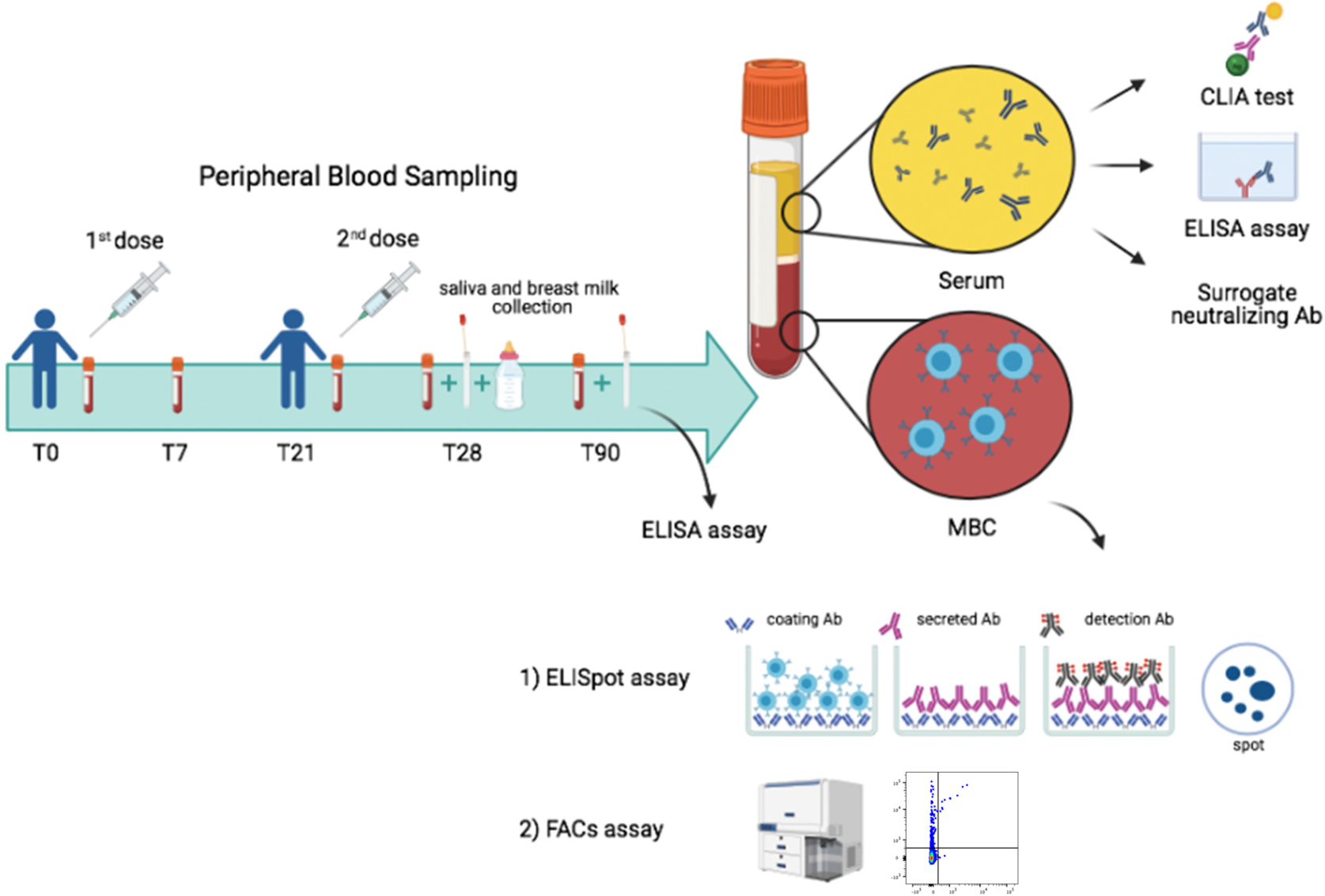
Study design. Design of our vaccine study indicating timeline and experimental plan.

Specific antibodies were measured in the serum, saliva and breast milk (Fig. 1). MBCs were identified by two different methods. By ELISpot, after polyclonal stimulation with CpG, we detected MBCs, based on their ability to secrete antibodies that bind the vaccine target Trimeric Spike on coated plates. We also identified MBCs *ex-vivo* by flow-cytometry, thanks to the availability of biotin-labelled Trimeric Spike coupled to a very high brightness fluorescence dye (PE) and the same biotin-labelled trimeric Spike coupled to a moderate brightness fluorescent dye (BUV395)^14^. We were able to distinguish MBCs with low (PE single positive, S+) or high affinity (PE-BUV395 double positive, S++) for Trimeric Spike.

### Serum antibodies

Serum antibodies are the most commonly used surrogate biomarkers of vaccine efficacy. We measured specific serum antibodies using different methods.

Anti-RBD antibodies of IgM isotype were measured by ELISA. We found that anti-RBD IgM was present at low levels at T0 and T7 reflecting the presence of natural^11, 15^ or cross-reactive antibodies^16, 17^. The increase at T21 and T28 is the effect of the vaccination. At T90 anti-RBD IgM slightly declined (Fig. 2a).

**Fig. 2.**
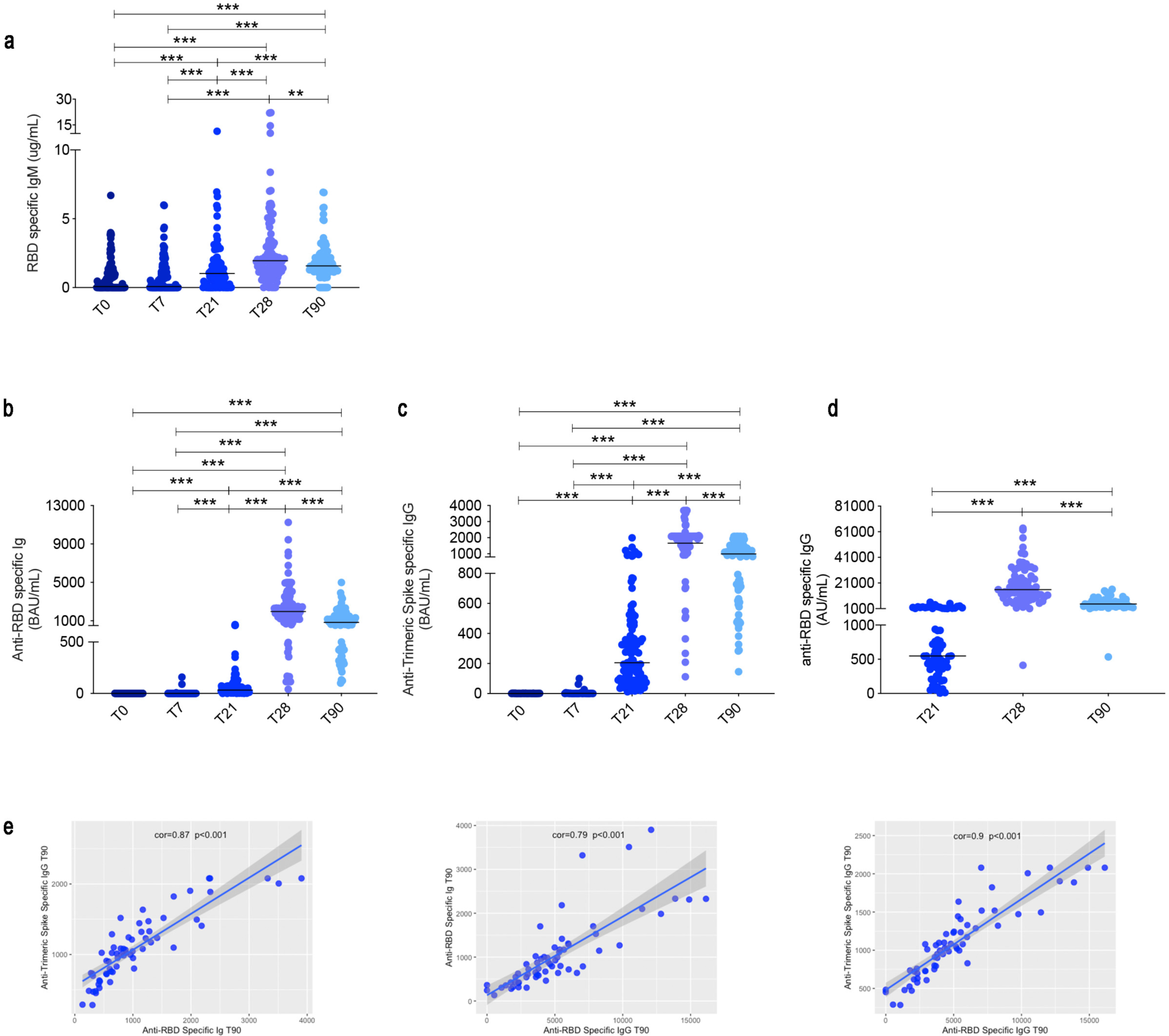
SARS-CoV-2 Antibody Responses. **(a)** Anti-RBD IgM was measured by an *in-house* ELISA. (**b)** Anti-RBD and **(c)** Anti-trimeric S titers were assessed by Elecsys® Anti-SARS-CoV-2 S assay (Roche, cut-off 0.8 U/mL) and Liaison SARS-CoV-2 trimeric S IgG Assay (Diasorin, cut-off 13 AU/mL) before vaccine administration (T0), at 7 (T7), 21 (T21) 28 (T28) and 90 (T90) days after the first dose (*n=108*). **(d)** Surrogate neutralization IgG titers at T21, T28 and T90 (*n=70*). (**e**) Scatter plots with a regression line of IgG Anti-trimeric S *vs* Ig anti-RBD; Ig anti-RBD *vs* IgG anti-RBD and IgG Anti-trimeric S *vs* IgG anti-RBD. On top of the graph we reported the Pearson’s correlation coefficient and the p-value from the correlation test.Medians are plotted and statistical significance was determined using Wilcoxon matched-pairs signed-rank test. *p < 0.05, **p < 0.01, ***p < 0.001.

Anti-RBD antibodies of all isotypes, evaluated by Elecsys®Anti-SARS-CoV-2-S (Roche, cut-off: 0.8 BAU/mL) were absent at T0 and T7, but already increased at T21. Their level, however, was much higher at T28 and decreased at T90 (Fig. 2b). The geometric mean at T21 was 29.13 BAU/mL and became 1736 BAU/mL at T28, demonstrating the importance of the booster dose. At T90 the geometric mean decreased to 819.8 BAU/mL. Similar results were obtained when we measured IgG against Trimeric Spike (DiaSorin, cut-off: 13 BAU/mL). The geometric mean values increased from 205.9 BAU/mL at T21 to 1660 BAU/mL at T28 and then decreased to 991.3 BAU/mL at T90 (Fig. 2c).

Thus, specific antibodies are already produced after the first vaccine dose. However, anti-RBD antibodies increased 60 times between T21 and T28, and anti-Trimeric Spike IgG raised 8 times after the second dose. Three months after the first dose anti-RBD antibodies and anti-Trimeric Spike IgG were reduced by half (Fig. 2b,c).

The anti-viral activity of IgG after the first and second dose and at T90 was measured by using, as surrogate virus neutralization assay, a specific chemiluminescence microparticle antibody assays (CMIA) on Abbott ARCHITECT® i2000sr^18^ that detects IgG antibodies specific for RBD. The geometric mean at day 21 was 559.3 AU/mL and increased 25 times at T28 (geometric mean 14129 AU/mL), thus confirming the importance of re-stimulation to obtain a significant amplification of the protective response (Fig. 2d). Surrogate-neutralization activity was reduced by 3.2 folds at T90 (geometric mean 4403; Fig. 2d).

The reduction of antibody levels three months after vaccination reflects the normal kinetics of the serological immune response^19^.

There was no correlation between antibody levels measured by the different tests at early times after vaccination (data not shown), whereas the correlation became significant at T90 (Fig. 2e). As the commercial test used here measure slightly different responses (all isotypes against RBD, IgG specific for either Trimeric Spike or RBD), the correlation at T90 may reflect the loss short-lived plasma cells released in the early stages of the immune reaction and the progressive selection of useful IgG antibodies against trimeric Spike and RBD.

### Memory B cell detected by ELISpot

ELISpot to detect B cells able to secrete Trimeric Spike-specific IgM, IgG and IgA antibodies was performed in 74 HCWs. We found that a high frequency of IgM MBCs was able to bind Trimeric Spike already at T0 and their number slightly increased 7 days after the first dose (T7) (Fig. 3a). This is an expected result, because our pool of IgM MBCs includes a wide range of cross-reactive specificities from which high affinity MBCs will be shaped and selected by the germinal center (GC) reaction^12^.

**Fig. 3.**
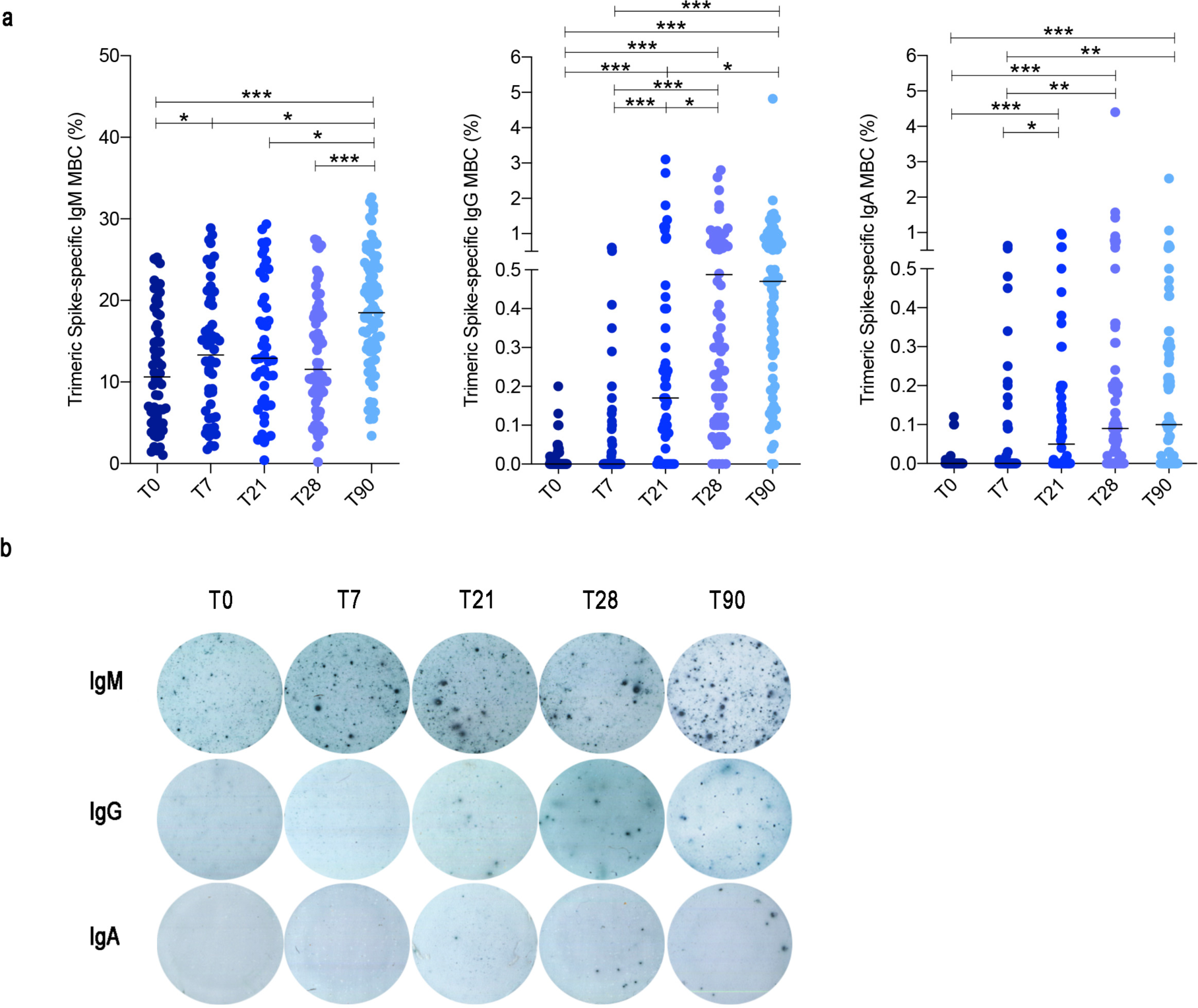
Antigen specific memory B cell responses after mRNA BNT162b2 vaccination in HCW. **(a)** Trimeric Spike-specific MBCs were detected by ELISpot following PBMC stimulation with CpG, IL-21 and IL-4 for 5 days. For each isotype, results are shown as percentage (%) of MBCs specific for Trimeric Spike over time (T0, T7, T21, T28 and T90). **(b)** Representative images of Trimeric Spike-specific MBCs detected by ELISpot. Medians are plotted and statistical significance was determined using Wilcoxon matched-pairs signed-rank test. *p < 0.05, **p < 0.01, ***p < 0.001.

In the GC, the mechanisms of somatic mutation and antigen-driven affinity-maturation improve the specificity of the antibodies and class-switching generates MBCs and plasma cells able to secrete switched antibodies that have the function of terminating infections and clearing the pathogen.

High specificity and affinity are the most important characteristics of protective MBCs, generated by the adaptive immune system in response to infection or vaccination.

In agreement with the timing of the GC reaction, IgG MBCs specific for Trimeric Spike significantly increased at T21 after the first dose and significantly more at T28, namely seven days after the second dose (Fig. 3a). Importantly, we found that 20.8% of the HCWs did not have IgG-producing MBCs cells in the peripheral blood at T21, following the first dose. Seven days after the second dose (T28), however, IgG-producing MBCs became detectable in the great majority of the HCWs (92.4%) (Fig. 3a,b), again demonstrating the importance of the booster administration.

IgA-secreting MBC were less frequently detected in the peripheral blood of vaccinated individuals. We found that 58.4% of the HCWs produced IgA MBCs 21 days after the first dose; seven days after the second dose (T28), IgA-producing MBCs were found in the peripheral blood of 71.3% of the individuals (Fig. 3a,b).

An important finding of our study is that, in contrast to the decline of specific antibody levels in the serum, MBCs of IgM, IgG and IgA isotypes remained stable three months after the first dose (Fig. 3a,b).

Overall, our results show that at all time points of analysis, about 10% (median) of IgM MBCs were able to secrete antibodies binding Trimeric Spike in ELISpot. Our data confirms the role of IgM MBCs as first-line defence^20^ and substrate^12^ for the generation of high-affinity switched MBCs. The adaptive immune response triggered by the vaccine needs time and is strongly selective^21^.

For this reason, IgG and IgA MBCs specific for Trimeric Spike significantly increase only at T21 and T28 and are rare. 0.17% of the IgG MBCs are specific for Trimeric Spike at T21 and 0.48% at T28. For IgA MBCs the corresponding medians are 0.05% at T21 and 0.09% at T28 (Fig. 3a). At T90 there was no further change of IgG and IgA MBCs. IgM MBCs instead significantly increased suggesting that the memory B cell pool maintains and expands the newly acquired specificity in its adaptable repertoire, represented by IgM MBCs^12^.

### Spike-specific memory B cells detection by flow-cytometry

MBCs can be detected in the peripheral blood of convalescent patients and immunized individuals using fluorescent Trimeric Spike and RBD^22^.

We chose to visualize biotin-labelled Trimeric Spike using two different streptavidin-fluorochromes, one with a very high brightness index (streptavidin-PE) and the other with a moderate one (streptavidin-BUV395). Our intention was to visualize low-affinity MBCs detectable with streptavidin–PE (S+) and high-affinity MBCs that would appear as double positive (PE-BUV395; S++). Indeed, in individuals convalescent from COVID-19 infection most MBCs have a high-affinity and are double positive for PE and BUV395 Trimeric Spike (S++) (Extended Data Fig. 1). S++ MBCs are absent in HCWs who have never experienced SARS-CoV-2 infection before and their development is induced by the vaccine.

Among Spike positive MBCs (S+ plus S++), we also detected RBD-specific cells with Streptavidin - FITC bound to biotin-RBD. After excluding aspecific binder B cells with streptavidin-PE-Cy7, we gated CD19+CD24+CD27+CD38-MBCs (Fig. 4a,b) and calculated the frequency of S+ and S++ MBCs and of RDB binder. We found that low affinity MBCs (S+) were already detectable at T0 (Fig. 4c,d) and were mostly of IgM isotype (Fig. 4e), as expected, because of the structure and function of the large repertoire from which we shape and select our specific MBCs and in agreement with our ELISpot results (Fig. 3a,b). S+ MBCs slightly increase at T7 and more significantly at T21 and T28 (Fig. 4d) remaining mostly of IgM isotype (Fig. 4e).

**Fig. 4.**
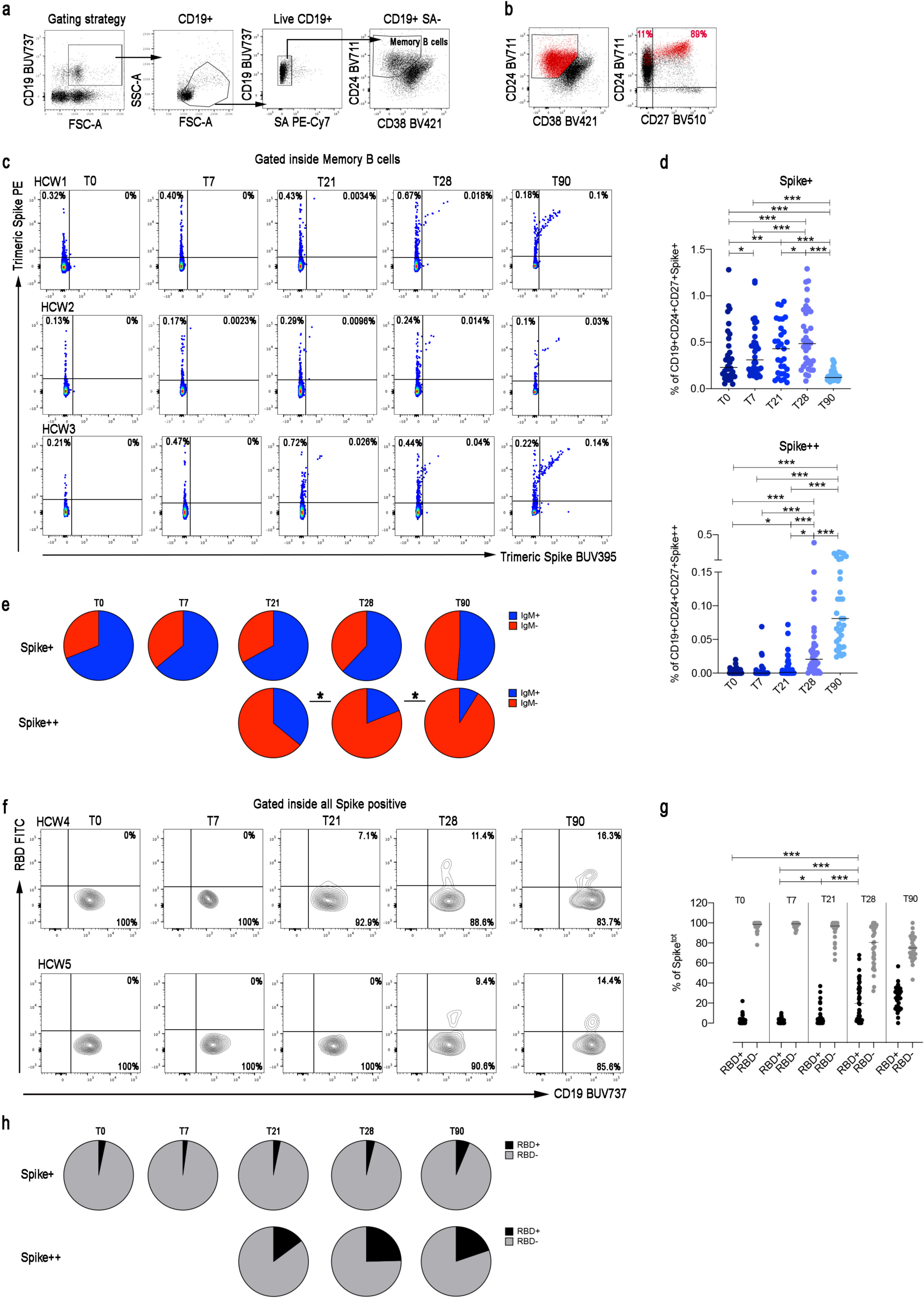
Trimeric-Spike specific memory B cells. **(a-b)** Gating strategy to identify S+ and S++ MBCs. S+ MBCs were identified as CD19+PECy7-CD24+CD27+CD38-Spike-PE+SpikeBUV395-; S++ MBCs were gated as CD19+PECy7-CD24+CD27+CD38-Spike-PE+SpikeBUV395+. **(c)** Flow cytometry plots in three representative HCWs showing staining patterns of SARS-CoV-2 antigen probes on MBCs during the follow-up. **(d)** Scatter plots depict the percentage of S+ and S++ inside MBCs (*n=34*). **(e)** Pie charts represent the percentage of IgM+ and IgM-S+ and S++ MBCs. **(f)** Identification of RBD+ cells inside total Spike positive (S+ plus S++) MBCs in two representative HCWs. **(g)** Scatter plot shows the percentage of RBD+ and RBD-Spike specific MBCs at the different time points of the analysis (*n=34*). **(h)** Pie charts show the average percentages of RBD+ and RBD-among MBCs specific for Trimeric Spike (S+ plus S++). Medians are plotted and statistical significance was determined using Wilcoxon matched-pairs signed-rank test or Chi-Square test. *p < 0.05, **p < 0.01, ***p < 0.001.

Double positive Trimeric Spike-specific MBCs (S++) significantly increased at T21 and even more at T28, seven days after the second dose (Fig. 4c,d). Most of them expressed switched isotypes (IgM-) (Fig. 4e).

At T90 we found that S+ MBCs significantly declined, whereas the frequency of S++ MBCs increased 4 fold (Fig. 4c,d). These results reflect the ongoing maturation of the germinal center response leading to the progressive selection of MBCs that optimally bind the antigen and are mostly of switched isotypes. At T90 the majority of S+ MBCs were still of IgM isotype (Fig. 4e).

Among all MBCs able to bind Trimeric Spike (S+ plus S++), we identified RBD+ cells that represent a minority of the MBCs generated by vaccination. RBD+ MBCs significantly increased at T21 and even more at T28. Their frequency remained stable at T90 (Fig. 4f,g). RBD+ MBCs were almost only found in the S++ MBC pool (Fig. 4h), indicating the need of the germinal center mutation and selection mechanisms to generate this new specificity.

### IgA is not produced at mucosal sites after vaccination with BNT162b2

The production of IgA is of great interest, because IgA is the main immunoglobulin for protection at mucosal sites, including the upper airways, the site of SARS-CoV-2 entry.

We measured Spike-specific IgA in the serum of immunized HCWs. We found that after the first dose 22% of the HCWs had not produced Spike–specific IgA, but all of them had specific IgA at T28 in the serum. The level of IgA strongly decreased at T90 (geometric mean 12.75 at T28 and 2.6 at T90) indicating that most of the IgA plasma cells generated in response to the vaccine were short-lived (Fig. 5a).

**Fig. 5.**
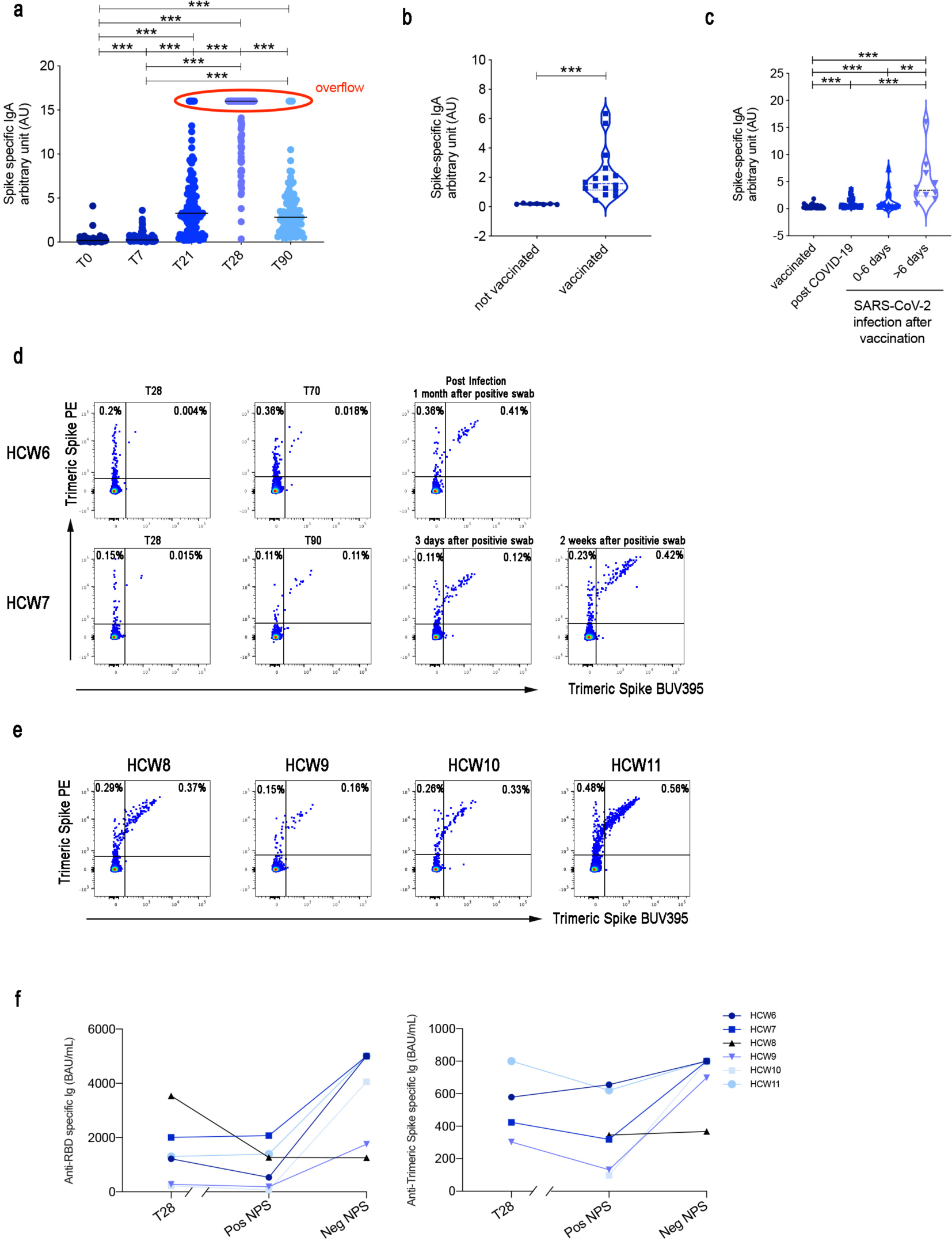
SARS-CoV-2 IgA Antibody Response. **(a)** Spike specific IgA antibodies were detected by ELISA at different time points in the serum of all HCWs included in the study (*n=108*). Overflow values are indicated by a red oval. (**b**) Spike specific IgA antibodies were measured in the breast milk of 7 controls and 16 HCWs mothers vaccinated after the delivery. **(c)** Spike specific IgA antibodies were measured in the saliva of 43 representative HCWs with the highest serum specific IgA levels, in 39 HCWs who had recovered from COVID-19, and in 6 HCWs infected after two vaccine doses. In the last group specific salivary IgA was detected at the indicated days after the first positive nasopharyngeal swab. (**d-e**) Flow cytometry plots of 6 HCWs infected after two vaccine doses showing the staining patterns of SARS-CoV-2 antigen probes on MBCs. (**f**) Anti-RBD and Anti-trimeric S titers were measured in the 6 HCWs who had a positive nasopharyngeal swab after vaccination at T28, at the time of positive nasopharyngeal swab (Pos NPS) and when they return to work (Neg NPS). The interrupted x-axis indicates that after T28, the time of the first positive NPS was variable (between day 60 and day 110 after first dose) and longer compared to the time between the positive and negative NPS (2 to 20 days). Medians are plotted and statistical significance was determined using Wilcoxon matched-pairs signed-rank test or unpaired Mann-Whitney t-test. *p < 0.05, **p < 0.01, ***p < 0.001.

Antibodies can reach mucosal sites by transudation from the serum or be locally produced after MBCs have migrated to mucosal sites and have become plasma cells. The latter mechanism is indispensable for the secretion of IgA into breast milk^23^. Pro-inflammatory cytokines and chemokines are physiologically increased in the lactating breast^24, 25^ and switched MBCs primed to secrete antibodies can be found in maternal milk. Sixteen HCWs were vaccinated after pregnancy, during lactation (Table 3). Seven days after the second vaccine dose, all of them had detectable IgA specific for trimeric Spike in breast milk (Fig. 5b) confirming the ability of vaccine-induced MBCs to home to the inflammatory environment of the lactating mammary gland and locally produce IgA.

**Table 2:** Study cohort used in the different assays.

| Number of patients | Assay | Total patients followed over time |
| --- | --- | --- |
|  | ELISpot | 74 |
|  | FACs | 34 |
|  | Surrogate Neutralization | 70 |
|  | ELISA | 108 |

**Table 3:**
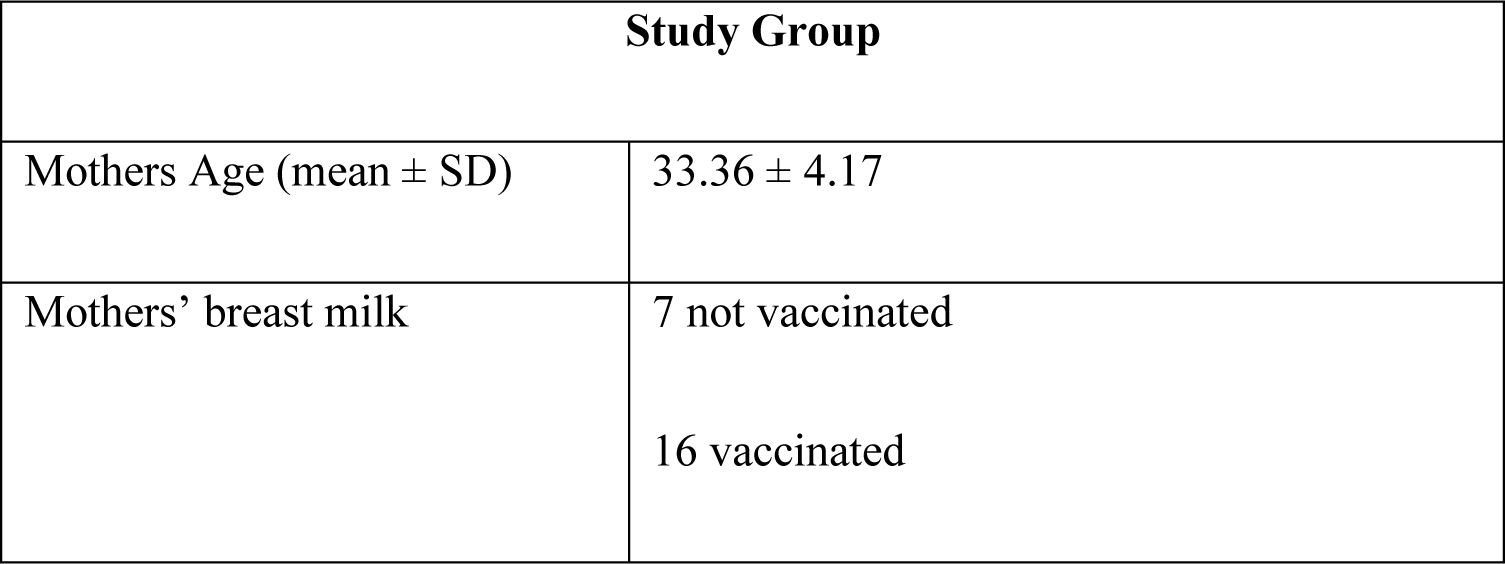
Characteristics of enrolled mothers.

| Study Group |  |
| --- | --- |
| Mothers Age (mean ± SD) | 33.36 ± 4.17 |
| Mothers' breast milk | 7 not vaccinated<br>16 vaccinated |

Saliva can be considered a proxy of mucosal immunity containing secretory IgA produced by local plasma cells^26^. In order to investigate whether serum IgA is associated with mucosal protection after vaccination, we measured IgA in the saliva of 43 HCWs that had the highest IgA values in the serum at day 28.

We found that two to three weeks after the second dose, IgA was undetectable in the saliva of all vaccinated individuals tested (Fig. 5c), demonstrating that the vaccination does not result in the presence of significant levels of IgA in the oral mucosa. Specific IgA was detectable in the saliva of 39 HCWs previously infected with COVID-19 (Fig. 5c), indicating that, as published before, the disease induce mucosal immunity^27^.

Twenty-one of the 3511 vaccinated HCWs of our Institute had a positive swab after completion of the two-dose vaccine cycle. All of them were related to familiar clusters. All the infected HCWs had responded to the vaccine with serum antibody production before the positive nasopharyngeal swab. The infection was either asymptomatic or mild, without signs of pneumonia. We studied the immune response of 6 HCWs infected after vaccination (Table 4). Salivary IgA became detectable 0-6 days after the first positive swab and increased further after 6 days from the infection (Fig. 5c). Specific IgA in the saliva was 10 times lower than IgA in the serum.

**Table 4:** Characteristics of HCWs infected after two doses of vaccine.

|  |  |
| --- | --- |
| **HCWs Age (mean ± SD)** | 45.6 ± 6.5 |
| **Sex** | 66.7 % F; 33.3% M |
| **SARS-CoV-2 infection** | 33.3% asymptomatic; 50% paucisymptomatic;<br>16.7% mild<br>5 |
| **SARS-CoV-2 variant** | 66.6% P.1; 16.7% B.1.1.7 ; 16.7% n.a. |

Two of the HCWs who had a positive swab after the second vaccine dose were included in our MBC study. From the first one we compared the samples obtained at the T28, at T70, and at T118 when the HCW returned to work. During the infection, caused by a virus variant (P.1), the initial symptoms were mild, with fever, cough and muscle pain. Most of the MBCs were of low affinity (S+) at T28 and T70. S++ MBCs were increased 23 times in the last sample (Fig. 5d).

In contrast, the second HCW had both S+ and S++ MBCs at T28 and T90. A nasopharyngeal swab resulted positive 20 days after T90 and the MBCs pool remained stable in the next three days, and strongly increased 2 weeks later (Fig. 5d). The infection was asymptomatic in this case.

Of 4 HCWs that were not included in the initial study and had a positive swab between T60 and T80, we obtained a sample when they returned to work after a paucisymptomatic disease. Their S++ were in higher frequency compared to the HCWs in the initial study group (median=0.08%, in Fig. 4d) confirming that MBCs generated by the vaccine increase in response to the virus (Fig. 5e). A further evidence of the function of MBCs is the increase of serum specific IgG after the positive swab in 5 out of 6 HCWs (Fig. 5f). HCW8 was found to have a positive swab during a routine control, in the absence of symptoms. The nasopharyngeal swab became negative after two days. High levels of salivary IgA (overflow) were detected at day 7 (the only sample available). Retrospectively we found that HCW8 had antibodies against the viral nucleoprotein already after the complete vaccine cycle, in February. We hypothesize that a previous asymptomatic and undetected infection had resulted in a powerful mucosal protection able to eliminate the virus in two days and prevent the systemic response.

## Discussion

The aim of vaccination is to generate immune memory, namely the pool of cells, including high-affinity MBCs and long-lived plasma cells with their antibodies that are able to prevent reinfection. Specificity and rapidity of action are the indispensable properties of protective immune memory. Differently from memory T cells, MBCs have the ability to improve their specificity by repeated steps of somatic mutation and selection in the GC. MBCs with high affinity for Trimeric Spike produce antibodies able to neutralize the virus. Monoclonal antibodies obtained after cloning from these MBCs can be used for cure and prevention of SARS-CoV-2 infection^28^.

Few days after a vaccine cycle, specific antibodies peak in the serum thanks to the activity of short and long-lived plasma cells^9^. The decay of short-lived plasma cells^9^ causes the reduction of serum antibodies. As no vaccination before has been under such a constant and severe scrutiny by the public and media as this against SARS-CoV-2, the physiological drop of serum titers is seen as a sign of vaccine failure and loss of protection. In response to this anxiety, the necessity of further vaccine booster doses has been suggested.

Although three months after the first dose serum specific antibody levels decline, we found that MBCs generated in response to vaccination continue to improve their specificity and increase in numbers.

By ELISpot we show that the immune response to the vaccine generates specific MBCs expressing switched isotypes at T28, seven days after the second dose in most individuals, whereas at T21 MBCs of switched isotypes are still undetectable in 20.8% and in 41.6% of the immunized subjects, for IgG and IgA respectively (Fig. 3). The abundance of IgM MBCs binding Trimeric Spike seen by ELISpot at all time-points contrasts with the appearance of IgG and IgA secreting cells only after one and two doses of vaccine. Our results can be explained considering that the ELISpot also detects low affinity pentameric IgM and MBCs expressing and secreting this type of antibodies that play an important role in the functional organization of the MBCs pool. A large group of MBCs able to bind the antigen with low affinity is important as the first step of defence. Only a few among these cells will be recruited in the GC reaction. The stringency of selection in the GC, explains why very few of the IgM MBCs that bind Trimeric Spike in ELISpot at all time-points (median 10%) become IgG (median 0.48%) or IgA (median 0.09%) secreting cells at T28. At T90 the frequency of IgG and IgA secreting MBCs remained stable.

In agreement with the ELISpot data, MBCs with low affinity for Trimeric Spike (S+), mostly of IgM isotype are present at all time-points (Fig. 4c and d), whereas high affinity MBCs become detectable at T21 and significantly increase further at T28. They are mostly of switched isotypes (Fig. 4c and d). At T90 the frequency of high-affinity MBCs is further increased, suggesting that the germinal center reaction continues its rounds of selection of the best specificity for months after the second vaccine dose. Low-affinity MBCs remain detectable and are still mostly of IgM isotype. MBCs able to bind RBD are about 20% of the S++ MBCs at day T28 and T90. Thus, a fraction of the response generated by immunization against trimeric Spike is directed against RBD. This is important to secure the anti-viral ability of the antibodies, although the specificity of neutralizing antibodies is not exclusively limited to the RBD domain^29^.

IgM MBCs with low-affinity for Trimeric Spike (detected by FACS as S+ and also measured in ELISpot) are present at all time points. At T0 they correspond to natural/innate MBCs^11^ or to MBCs involved in the response to previously encountered members of the coronavirus family^16^. IgM MBCs increasing and persisting after vaccination will be useful because they can be re-modelled in the GC in response to viral variants^12^.

The first obstacle posed by the immune system to mucosal infection is secretory IgA^30^. SIgA plays a fundamental role in the protection from respiratory viruses by blocking their attachment to epithelial cells^27, 31, 32^. It has been shown that adult patients with COVID-19 have high levels of specific and neutralizing SIgA in the saliva^27^. In order to prevent viral invasion of the epithelial cells of the upper respiratory tract, we need mucosal immunity and secretory IgA^33^, as demonstrated by the protective potency of maternal milk^34, 35^ and salivary IgA in children^36^. The absence of IgA in the saliva of vaccinated HCWs suggests that, in strict terms, the vaccine is not sterilising, because it is unable to generate preventive mucosal immunity.

The lack of direct mucosal protection explains why vaccinated individuals can have a positive nasopharyngeal swab. In most cases, however, the infection remains asymptomatic or mild. Part of their defence may be due to Trimeric Spike-specific IgG and IgA antibodies exudated to the tissues. An important role may also be played by MBCs that migrate to the infected areas. We show that MBCs generated by the vaccine are able to migrate to mucosal tissues and locally produce IgA antibodies in two different situations, both characterized by the production of attracting inflammatory cytokines. First we show that Spike-specific IgA can be detected in the breast milk of women vaccinated after pregnancy (Fig. 5b). Secondly, we found that HCWs with a positive swab after the complete cycle of vaccination, produce salivary IgA 2-4 days after the first positive swab (Fig. 5c). suggests that vaccine-induced MBCs have performed their job by rapidly migrating to the site of viral invasion. IgA in the saliva is of the secretory type, dimeric and bound to the secretory component. Dimeric IgA has a much more potent neutralization ability than serum monomeric IgA^37^. Thanks to the combined action of antibodies and MBCs (Fig. 5c-e) the evolution of the disease is immediately blocked. For this reason, the viral load in vaccinated individuals who present a positive nasopharyngeal swab can be so low to be unable to transmit the disease to others^38^.

The absence of cases of re-infection in the 299 HCWs, who experienced COVID-19 before vaccination and have detectable levels of salivary IgA (Fig. 5c), further highlights the importance of mucosal protection against SARS-CoV-2.

Mucosal vaccines against COVID-19 are in development^39^ and may more effectively prevent infection and abrogate viral circulation.

Real-world efficacy of one and two doses of RNA vaccines has been just reported by a CDC study. Among HCWs, one dose had 80% efficacy whereas two doses resulted in 90% efficacy^4^. In a different setting, in individuals over 60, after the first dose of BNT162b2 vaccine there was a significant decrease, but only of about 33% in the rate of positive tests for the coronavirus. The efficacy increased to 60% after two doses^5^. Many factors contribute to vaccine efficacy including age and viral circulation, but from the immunological point of view two doses are more protective than one. The real-world efficacy of one vaccine dose, demonstrated after day 14 post-immunization, can be explained by the performance of low-affinity MBCs and antibodies able to prevent severe COVID-19. Nevertheless, our data show that two doses of vaccine ensure a more protective and possibly long-lived immunological memory^40^.

We show that the strategy of delaying the second dose, recently proposed in Europe under the pressure of the spreading pandemic and the scarcity of vaccine doses may result in an imperfect immunological memory lacking the strength (high-affinity MBCs and antibodies) to rapidly and completely clear the virus. The persistence of replicating viruses in a partially protected patient might lead to the evolution and selection of viral variants. Variants rapidly spread and weaken the protective potentials of the vaccines that we already have and of those that are still in development.

The robustness of vaccine-induced protection does not depend only on the amount of serum antibodies, but mostly on the persistence of MBCs able to migrate to the site of infection, locally produce antibodies and remodel in response to viral variants. IgM MBCs that have acquired the specificity for Trimeric Spike may be the substrate for further modifications and adaptation to viral variants, thus ensuring a rapid response. In agreement with our hypothesis, vaccinated HCWs who had a positive nasopharyngeal swab experienced a pauci or asymptomatic infection by Brazilian (P.1) or UK (B.1.1.7) VoC. Further studies are necessary to investigate the duration and resilience of the immune memory induced by the available vaccines.

For all these reasons we should not worry about the antibody decline in the serum, observed a few months after vaccination or natural infection. Our MBCs are our most important defense weapon that ensure local and systemic protection after re-encounter with the antigen. In conclusion, as any other vaccine produced before, also RNA-vaccines appear to be able to generate a typical immunological memory that is predictably stable and durable.

## Data Availability

All data are available in the main text or the supplementary materials.

## Acknowledgments

We thank the study participants for their dedication to this project. Marco Scarsella and Ezio Giorda for the assistance with the FACs acquisition. Sandra Martino for technical coordination, Patrizia Palomba, Simona Cascioli, Priscilla Caputi, Ilaria Scibetta, Gloria Deriu, Matteo D’Agostini and Silvia Piras for the samples collection. We acknowledge the nurse staff for their support for the vaccination campaign and all the HCWs that have been enrolled for the study.

## Funding

This work was founded by:

Italian Ministry of Health RF2013-02358960 grant

Italian Ministry of Health COVID-2020-12371817 grant

Ricerca Corrente 2021 “5 per mille”

## Author contributions

Conceptualization: RC, EPM, CR, MRV, ST, AFS, CFP, SZ, TC

Methodology: EPM, ST, AFS, CAlb, PR, MS, GR, EP, CR, LP, CC, SM, MRV, RB, DG, NR, GS, MCDA, CAlt, LC, AR, NC, DA, MA

Data interpretation: RC, EPM, ST, AFS, CR, MRV

Funding acquisition: RC, CAg, FL, CFP, SZ

Project administration: RC, CFP, SZ

Supervision: RC, CFP, SZ

Writing – original draft: RC, EPM, ST, AFS

Writing – review & editing: RC, EPM, ST, AFS, FL, CFP, SZ, NM, MRV, CR

## Competing interests

Authors declare that they have no competing interests.

## Data and materials availability

All data are available in the main text or the supplementary materials.

## Online materials and methods

### Ethical Approval

Ethical approval was obtained from the Ethics Committee at OPBG, Bambino Gesù Children Hospital. According to the guidelines on Italian observational studies as established by the Italian legislation about the obligatory occupational surveillance and privacy management; Health Care Wokers (HCWs) confidentiality was safeguarded, and informed consent was obtained from all the participants. The study was performed in accordance with the Good Clinical Practice guidelines, the International Conference on Harmonization guidelines, and the most recent version of the Declaration of Helsinki.

### Human subjects

We analysed peripheral blood from 108 HCWs of the Bambino Gesù Children Hospital (Table 1 and 2). We included in the study all HCWs that were vaccinated by two injections of BNT162b2, 30 μg, 21 days apart.

Blood and serum samples were collected at defined intervals after vaccination. Samples were available before first dose (T0), and at 7 (T7), 21 (T21), 28 (T28) ant 90 (T90) days after the first dose. All HCWs had a negative SARS CoV-2 status by molecular (Allplex2019-ncov, Seegene) and antibody assays (Elecsys®Anti-N, Roche).

At the time of blood sampling, none of the subjects had an acute infection or were taking any medication known to alter immune function (such as steroids or statins).

Given the limiting amount of blood available for the study, we randomly selected 74 HCWs samples for the ELISpot and 34 for FACs (Table 2). Serology was performed in all samples and surrogate neutralization in 70 samples and only at T21, T28 and T90 (Table 2).

Breast milk was obtained from 23 HCWs, of which 16 were vaccinated during lactation (Table 3).

Saliva samples were collected by aspirating with a 1 mL syringe from the oral cavity at least 30 minutes after after eating, drinking, smoking, or chewing gum. Saliva was immediately transfered to an eppendorf tube and frozen at -20°C until use.

### Cell isolation and cryopreservation

Heparinized PBMCs were isolated by Ficoll Paque™ Plus 206 (Amersham PharmaciaBiotech) density-gradient centrifugation and immediately frozen and stored in liquid nitrogen until use. The freezing medium contained 90% Fetal Bovine Serum (FBS) and 10% DMSO.

### Polyclonal memory B cell stimulation

As we published before ^41^ in order to induce antibody production from MBCs, we polyclonally stimulated PBMCs with CpG. Briefly, PBMCs were cultured in complete medium at a concentration of 1×10^6^ cells/mL. Complete medium was prepared as follows: RPMI-1640 (Euroclone), 10% heat inactivated HyClone FBS (Hyclone Laboratories), 1mM L-Glutamine (GIBCO BRL); 1× Penicillin/Streptomycin 100× (Euroclone), 1% sodium pyruvate (GIBCO BRL). Cells were stimulated for 5 days with 0.35µM TLR9 agonist CpG-B ODN2006 (Hycult Biotech) plus 20 ng/mL rhIL-21 (Milteny) and 20 ng/mL rhIL-4 (Milteny).

### B cell ELISpot

The B cell (enzyme-linked immunospot) ELISpot assay was performed with 5 days stimulated-PBMCs for the detection of MBCs.

96-well Multiscreen filter plates (Millipore) were pre-wetted with 35% ethanol for ≤ 1 min and washed with ddH_2_O (5 x 200 µl/well). Wells were coated with 50 µl PBS containing either isotype-specific AffiniPure F(ab’)_2_ Fragment Goat anti-Human antibody (Jackson ImmunoResearch Laboratories) (anti-IgM final concentration 2.5 µg/mL, anti-IgG 15 µg/mL, anti-IgA 10 µg/mL); or Trimeric Spike at a final concentration of 1 µg/ml to detect specific responses. The plates were then incubated overnight (ON) at 4°C. After washing with PBS (5 x 200 µl/well) and blocking with PBS + 1% BSA + 5% Sucrose (5 x 200 µl/well) for 2 h at RT, plates were filled with sterile RPMI + 10% FBS (100 µl/well) for 1 h at 37°C.

After 5 days of stimulation (as previously described), cells were harvested and washed once with RPMI before counting. Different concentration of PBMCs were used depending on the following conditions: total IgM, IgG and IgA: 1.5 x 104 PBMCs/well; specific IgM: 5 x 104 PBMCs/well; specific IgG and IgA: 1-5 x 105 PBMCs/well. Four steps of two-fold serial titrations were performed for each condition. Plates were incubated ON at 37°C. Cells were discarded and after washing with PBS + 0,05% Tween (5 x 200 µl/well), plates were incubated for 1 h at 37 °C with peroxidase-conjugated goat anti-human IgM, IgG or IgA antibodies (Jackson ImmunoResearch Laboratories) diluted in PBS + 0,05% Tween + 1% BSA. The plates were washed with PBS + 0,05% Tween (5 x 200 µl/well) before TMB substrate (Mabtech) addition and incubated for 4 min at RT in the dark. After stopping the reaction by rinsing with ddH_2_O (5 x 200 µl/well), plates were left to dry ON at 4 °C.

All plates were analyzed using an automated AELVIS ELISpot reader (A.EL.VIS GmbH, Germany). Total IgM, IgG and IgA were used as positive control for each subject at each time point. Samples were excluded if they generated low total Ig responses. For statistical analysis, the frequency of antigen specific MBCs was reported as percentage (%) of total MBCs producing antibodies of IgM, IgG or IgA isotype in our culture conditions^42^.

### ELISA for specific IgA and IgM detection

A semi-quantitative *in vitro* determination of human IgA antibodies against the SARS-CoV-2 was evaluated on serum, saliva and breast milk samples by using the Anti-SARS-CoV-2 ELISA (Euroimmun), according to the manufacturer’s instructions. Values were then normalized for comparison with a calibrator. Results were evaluated by calculating the ratio between the extinction of samples and the extinction of the calibrator. The ratio interpretation was as follows: <0.8 = negative, ≥0.8 to <1.1 = borderline, ≥1.1 = positive^43^.

To detect IgM anti RBD we developed an in-house ELISA. 96-well plates (Corning) were coated for 1 h at 37°C with 1 μg/mL of purified SARS-CoV-2 RBD protein (Sino Biological). After washing with PBS 1×/0.05% Tween and blocking with PBS 1×/1% BSA, plates were incubated for 1 h at 37°C with diluted sera (1:100). After washing again, plates were incubated for 1 h at 37°C with peroxidase-conjugated goat anti-human IgM antibody (Jacksons ImmunoResearch Laboratories). The assay was developed with o-phenylen-diamine tablets (Sigma-Aldrich) as a chromogen substrate. Absorbance at 450 nm was measured, and IgM concentrations were calculated by interpolation from the standard curve based on serial dilutions of monoclonal human IgM antibody against SARS-CoV-2 Spike-RBD (Invivogen)^43^.

### Quantitative determination of anti-N, anti-S, Trimeric Spike and RBD antibodies

Serum samples were tested by two different methods and analytical platforms. Qualitative detection of antibodies direct against the nucleocapsid (N) protein and semi-quantitative detection of total antibodies (IgA, IgM, IgG) directed against the RBD of the virus Spike (S) protein of SARSxxxxx-CoV-2 were tested by an electro-chemiluminescence sandwich immunoassay (ECLIA), using Elecsys-anti SARS-CoV-2 and Elecsys-anti SARS-CoV-2 S (Roche Diagnostics) test on a Cobas e801 analyzer following the manufacturer’s instructions. Results for anti-N antibodies are expressed as “present” or “absent” on the basis of a cut-off index (COI) COI ≥ 1.0 and COI < 1.0, respectively. Detection and quantification were automatically calculated for each sample in U/mL, equivalent to the Binding Arbitrary Unit (BAU)/mL of the first WHO International Standard for anti-SARS-CoV-2 immunoglobulins. Titer was interpreted as absent when < 0.8 U/mL (< 0.8 BAU/mL) and as present when ≥ 0.80 U/mL (≥ 0.8 BAU/mL). When antibodies titer was higher than 250 U/mL (250 BAU/mL), the instrument automatically executed a 20-fold dilution, ranging the upper limit of quantification to 5000 U/mL (5000 BAU/mL). The quantitative determination of anti-Trimeric spike protein specific IgG antibodies to SARS-CoV-2 was run on LiaisonXL platform by a new generation of chemiluminescence immunoassay (CLIA) TrimericS IgG assay (DiaSorin) with a quantification range between 4.81 BAU/mL and 2080 BAU/mL (dilution factor 1:20).

The SARS-CoV-2 IgG II kit, a chemiluminescence microparticle antibody assays detecting antibodies against the RBD of SARS-CoV-2, (CMIA, IgG antibody concentrations expressed as arbitrary units, AU/mL ≥ 50 are considered positive, ARCHITECT® i2000sr Abbott Diagnostics) was used according to manufacturer’s protocols.

### Detection of antigen-specific memory B cells

To detect SARS-CoV-2 specific B cells, biotinylated protein antigens were individually multimerized with fluorescently labelled streptavidin at 4°C for one hour ^22^. Biotinylated trimeric SARS CoV-2 Spike (S1) was purchased from R&D systems. RBD were generated in-house and biotinylation was performed using EZ-Link^TM^ Sulfo-NHS-LC-Biotin reaction kit (ThermoScientific) following the manufacturer’s standard protocol and dialyzed overnight against PBS. Biotinylated Spike was mixed with streptavidin BUV395 (BD) and streptavidin PE (BD) at 25:1 ratio and 20:1 ratio respectively. Biotinylated RBD was mixed with streptavidin FITC (BD) at 2.5:1 ratio. Streptavidin PE-Cy7 (BD) was used as a decoy probe to gate out SARS-CoV-2 non-specific streptavidin-binding B cells. The antigen probes prepared individually as above were then mixed in Brilliant Buffer (BD). ∼5x10^6^ previously frozen PBMC samples were prepared and stained with 85μL antigen probe cocktail containing 100ng Spike per probe (total 200ng), 27.5ng RBD and 20ng SA-PE-Cy7 at 4°C for 30min to ensure maximal staining quality before surface staining with antibodies as listed in table S1 was performed in Brilliant Buffer at 4°C for 30min. SARS-CoV-2 specific memory B cells were defined as CD19+CD24+CD27+CD38-Spike+ (S+) or CD19+CD24+CD27+CD38-Spike++ (S++).

Stained PBMC samples were acquired on FACS LSRFortessa (BD). At least 4x10^6^ cells were acquired and analyzed using FlowJo10.7.1 (BD). Phenotype analysis of antigen-specific B cells was performed only in subjects with at least 10 cells detected in the respective antigen-specific memory B cell gate.

### SARS-CoV-2 Variant determination

SARS-CoV-2 variant determination was performed by single-nucleotide-polymorphism (SNP) detection approach. In particular the CE approved Real Time PCR COVID-19 Variant Catcher (Clonit Srl, Milan, Italy, CE IVD), able to detect the spike 69-70del, E484K and N501Y, was used to discriminate between the B.1.1.7 and the B.1.351/P.1 strains. Briefly, viral RNA was extracted, retrotranscribed and amplified according to manufacturer’s protocol. Real time PCR was performed on CFX96 (Bio Rad Laboratories). In order to further discriminate between B.1.351 and P.1, the presence of SNP H655Y and A701V were further investigated by means of home-made sanger sequencing protocol. In brief, spike region encompassing amino acids 605-900 was retrotranscribed, amplified and sequenced by using two different sequence-specific primers (FWD: 5’TACTTCTAACCAGGTTGCT3’; REV: 5’CTATAAGCCATTTGCATAGCA3’). Sequences were obtained thanks to the automated sequencer ABI-3130 (Applied Biosystems). Negative and positive controls were included in each run to monitor carry-over contamination and PCR efficiency.

### Statistical analysis

Values were compared by the non-parametric Kruskal-Wallis test and, if not significant, the Wilcoxon matched pair signed-rank test was used for the comparison between time points of each subject. Pairwise comparisons were evaluated by the Mann-Whitney U test nonparametric test. The Chi-square test with 2 × 2 contingency tables was used to compare pie charts. A p-values < 0.05 were considered statistically significant. Statistical analyses were performed with GraphPad Prism 8.0 (GraphPad Software).

The relationship between variables was studied using a correlation test based on the Pearson’s product moment correlation coefficient cor(x, y) and follows a t distribution with length(x)-2 degrees of freedom if the samples follow independent normal distributions. If there are at least 4 complete pairs of observation, an asymptotic confidence interval is given based on Fisher’s Z transform (/https://stat.ethz.ch/R-manual/R-devel/library/stats/html/cor.test.html).

**Extended Data Fig. 1.**
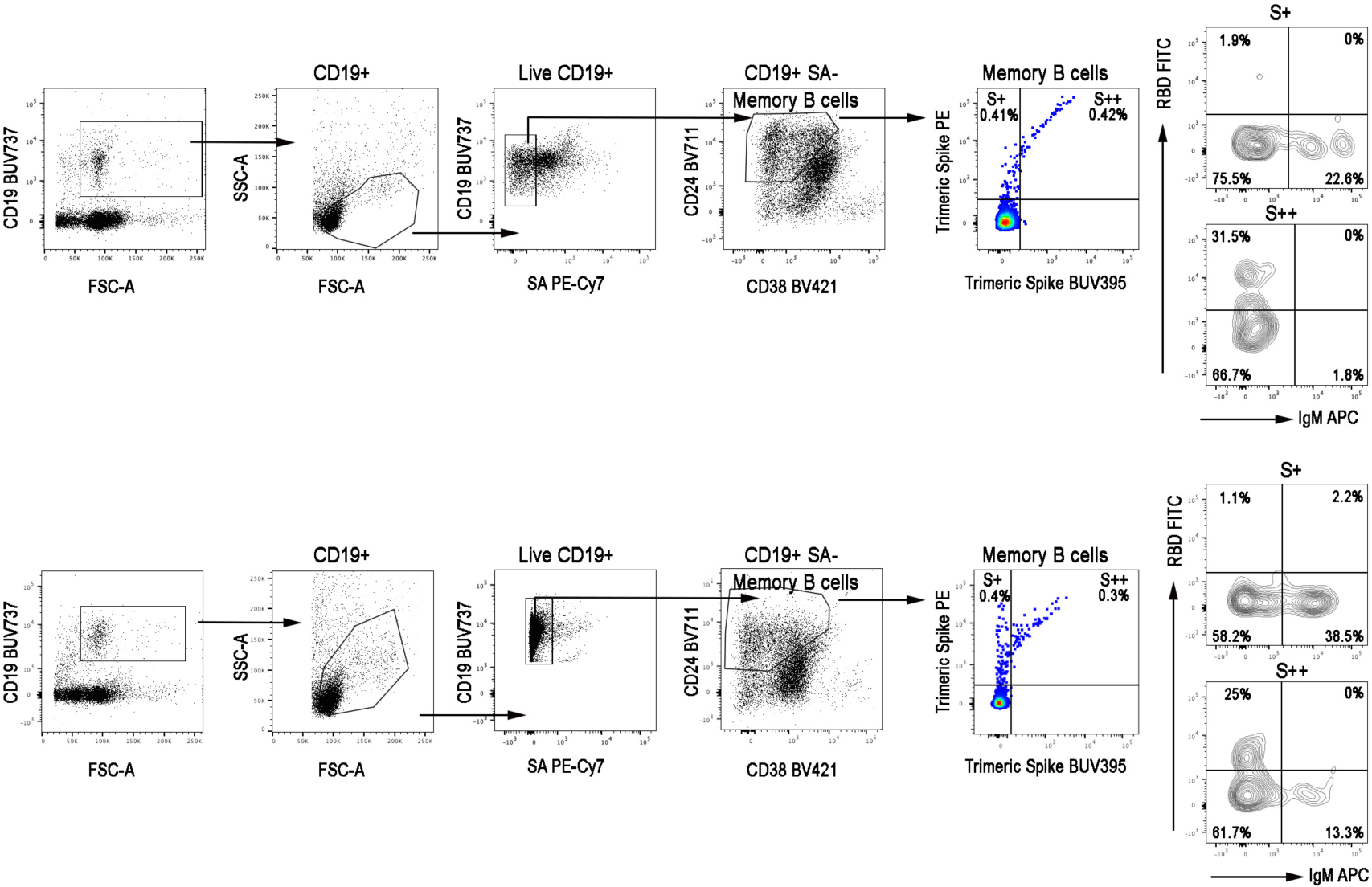
Trimeric-Spike specific memory B cells in convalescent COVID-19 patients. Gating strategy to identify S+ and S++ MBCs. S+ MBCs were identified as CD19+PECy7-CD24+CD27+CD38-Spike-PE+SpikeBUV395-; S++ MBCs were gated as CD19+PECy7-CD24+CD27+CD38-Spike-PE+SpikeBUV395+. Identification of RBD+ cells inside S+ and S++ MBCs in two representative convalescent patients.

**Supplementary Table 1:** Antibodies for staining.

|  | Clone | Catalog number |
| --- | --- | --- |
| CD19 BUV737 | SJ25C1 | 612757 |
| CD19 BV786 | SJ25C1 | 563325 |
| CD24 BV711 | ML5 | 563401 |
| CD27 BV510 | T-271 | 740167 |
| CD38 BV421 | HIT2 | 562444 |
| IgG BV650 | G18-145 | 740596 |
| IgM APC | Polyclonal | 709-136-073 |
| Streptavidin PE |  | 554061 |
| Streptavidin BUV395 |  | 564176 |
| Streptavidin FITC |  | 554060 |
| Streptavidin PE-Cy7 |  | 557598 |

## Notes

### Competing Interest Statement

The authors have declared no competing interest.

### Author Declarations

Ethical approval was obtained from the Ethics Committee at OPBG, Bambino Gesu Children Hospital. According to the guidelines on Italian observational studies as established by the Italian legislation about the obligatory occupational surveillance and privacy management; Health Care Wokers (HCWs) confidentiality was safeguarded, and informed consent was obtained from all the participants. The study was performed in accordance with the Good Clinical Practice guidelines, the International Conference on Harmonization guidelines, and the most recent version of the Declaration of Helsinki.

